# Prevalence of subpatent *Plasmodium falciparum* infections in regions with varying transmission intensities and implications for malaria elimination in Mainland Tanzania

**DOI:** 10.1101/2024.10.20.24315823

**Authors:** Misago D. Seth, Zachary R. Popkin-Hall, Rashid A. Madebe, Rule Budodo, Catherine Bakari, Beatus M. Lyimo, David Giesbrecht, Ramadhani Moshi, Ruth B. Mbwambo, Filbert Francis, Dativa Pereus, Doris Mbata, Daniel P. Challe, Salehe S. Mandai, Gervas A. Chacha, Angelina J. Kisambale, Daniel Mbwambo, Sijenunu Aaron, Abdallah Lusasi, Samwel Lazaro, Celine I. Mandara, Jeffrey A. Bailey, Jonathan J. Juliano, Julie R. Gutman, Deus S. Ishengoma

## Abstract

**Background:** Subpatent *Plasmodium falciparum* infections, defined as infections with parasitaemia density below the detection limit of routine malaria diagnostic tests, contribute to infectious reservoirs, sustain transmission, and cause the failure of elimination strategies in target areas. This study assessed the prevalence of and factors associated with subpatent *P. falciparum* infections in 14 regions of Mainland Tanzania with varying endemicity.

**Methods:** The study used samples randomly selected from RDT-negative dried blood spots (DBS) (n = 2,685/10,121) collected in 2021 at 100 health facilities across 10 regions of Mainland Tanzania, and four communities in four additional regions. The regions were selected from four transmission strata; high (five regions), moderate (three), low (three), and very low (three regions). DNA was extracted by Tween-Chelex method, and the *Pf18S* rRNA gene was amplified by quantitative polymerase chain reaction (qPCR). Logistic regression analysis was used to assess the associations between age groups, sex, fever status, and transmission strata with of subpatent infections status, while linear regression analysis was used to assess the association between these factors and subpatent parasite density.

**Results:** Of the selected samples, 525/2,685 (19.6%) were positive by qPCR for *P. falciparum*, and the positivity rates varied across different regions (range: 4.8 to 39.6%). Under-fives (aOR: 1.4, 95% CI 1.04-1.88; p<0.05) from health facilities had higher odds of subpatent infections compared to other groups, while those from community surveys (aOR: 0.33, 95% CI 0.15-0.72; p = 0.005) had lower odds. Participants from very low transmission stratum had significantly lower odds of subpatent infection compared to those from high transmission straum (aOR=0.53, 95% CI=0.37-0.78; p < 0.01). The log-transformed median parasite density (interquartile range) was 6.9 (5.8 - 8.5) parasites/µL, with significantly higher parasitaemia in the low transmission stratum compared to very low (11.4 vs 7.0 parasites/µL, p<0.001).

**Conclusion:** Even in very low transmission settings, the prevalence of subpatent infections was 13%, and in low transmission settings it was even higher at 29.4%, suggesting a substantial reservoir which is likely to be missed by routine malaria case management strategies. Thus, control and elimination programmes may benefit from adoption of more sensitive detection methods to ensure that a higher proportion of subpatent infections are detected.

## Introduction

Malaria remains a significant public health concern in many parts of the world, particularly in sub-Saharan Africa (SSA) [1]. In 2022, over half of the 608,000 malaria deaths occurred in four SSA countries: Nigeria (31%), the Democratic Republic of the Congo (12%), Niger (6%), and the United Republic of Tanzania (4%) [1]. In Tanzania, malaria is a leading cause of morbidity and mortality, with the entire population considered at risk of infection and 93% of the population living in areas where transmission occurs [2]. Malaria prevalence in Tanzania varies by region, with prevalence in under-fives ranging from 0 - 23.4% in 2022 [3]. Transmission intensity varies even within specific geographic areas [4–6]. This micro-epidemiology of malaria in Mainland Tanzania and the recent transition from holo/hyperendemic to hypoendemic transmission intensities need to be taken into account when planning for different malaria interventions, including those based on diagnostic and therapeutic methods.

Tanzania implements various control strategies as core interventions to control and eventually eliminate malaria; including malaria case management, vector control and chemoprevention. The vector control interventions include insecticide-treated mosquito nets (ITNs), indoor residual spraying, and larval source management, while case management methods are based on prompt diagnosis using rapid diagnostic tests (RDTs) and effective treatment with artemisinin-based combination therapies [4,7–9]. Currently, the only chemoprevention method used by the National Malaria Control Programme (NMCP) is intermittent preventive treatment in pregnancy using sulfadoxine-pyrimethamine (SP) [10]. Following recent significant progress in reducing cases and deaths, Tanzania has set an ambitious target to eliminate malaria as highlighted in the 2021-2025 National Malaria Strategic Plan (NMSP) [4]. In the current NMSP, enhanced surveillance, monitoring and evaluation, and response are highly prioritized to support the elimination efforts. In the current phase, which focuses on achieving malaria elimination, it is critical to ensure effective detection of malaria cases in very low transmission strata through passive, active, and reactive approaches. The efficacy of enhanced surveillance is dependent on the ability to detect as many infections as possible, including low-density subpatent infections, which are not detected by routine testing [11–13], and interfere with the efficacy of surveillance and control strategies.

Subpatent *Plasmodium falciparum* infections account for a substantial proportion of malaria infections (nearly 50% in some cases in asymptomatic individuals), and thus create a challenge for malaria control and elimination efforts, as such infections contribute to ongoing transmission but often go undiagnosed and untreated [14,15]. Due to the complex interaction between malaria immunity and transmission levels, no clear relationship between the prevalence of subpatent malaria infections and transmission intensity has been documented [14,16–18]. A number of studies have been conducted on the prevalence and significance of subpatent infections in areas with varying transmission intensities in Tanzania, and most of them have reported high prevalence in areas with low transmission intensities and low prevalence in areas with high transmission [19–25]. However, most of the studies report data that is limited in scope as each study covers a single geographically distinct area, making it difficult to obtain a nation-wide picture.

Subpatent infections have been shown to evade detection by routine diagnostic methods [14,26,27] and may act as reservoirs of and support ongoing transmission particularly in areas with very low transmission. Thus, in areas targeted for or implementing elimination strategies, this fraction of infection must be considered and addressed with specific, targeted interventions. This study utilized samples collected from surveys conducted in 2021 [28–30] to determine the prevalence and risk factors associated with *P. falciparum* subpatent infections in both asymptomatic and symptomatic individuals of all ages in 14 regions with varying malaria transmission intensity in Mainland Tanzania. The present study provides the first nation-wide scan of subpatent infections and forms a platform for future studies of such infections, with a focus on regions closer to the elimination targets.

## Materials and Methods

### Study sites and population

This study utilized data and samples collected from February to June 2021 in 14 regions spanning varying malaria transmission intensities as part of the project on molecular surveillance of malaria in Tanzania (MSMT) [28–30]. The study regions were selected among those located in four malaria burden strata based on the 2020 NMCP stratification; high (Geita, Kagera, Kigoma, Mtwara and Ruvuma), moderate (Mara, Tabora and Tanga), low (Dar es Salaam, Dodoma and Songwe) and very low transmission intensities (Kilimanjaro, Manyara and Njombe) (Fig. 1). In 10 regions (Dar es Salaam, Dodoma, Kagera, Kilimanjaro, Manyara, Mara, Mtwara, Njombe, Songwe and Tabora), cross-sectional health facility surveys were conducted in 10 health facilities per region, enrolling symptomatic patients with a history of fever in the preceding 48 hours or fever at presentation (axillary temperature >37.5°C) [28,29]. In four regions, participants were enrolled during community cross-sectional surveys (CSS) with samples collected irrespective of fever; in three of these (Kigoma, Ruvuma, and Tanga), individuals aged ≥6 months were enrolled, while in Geita, samples were collected from children aged 6 - 59 months only [30].

**Fig. 1:**
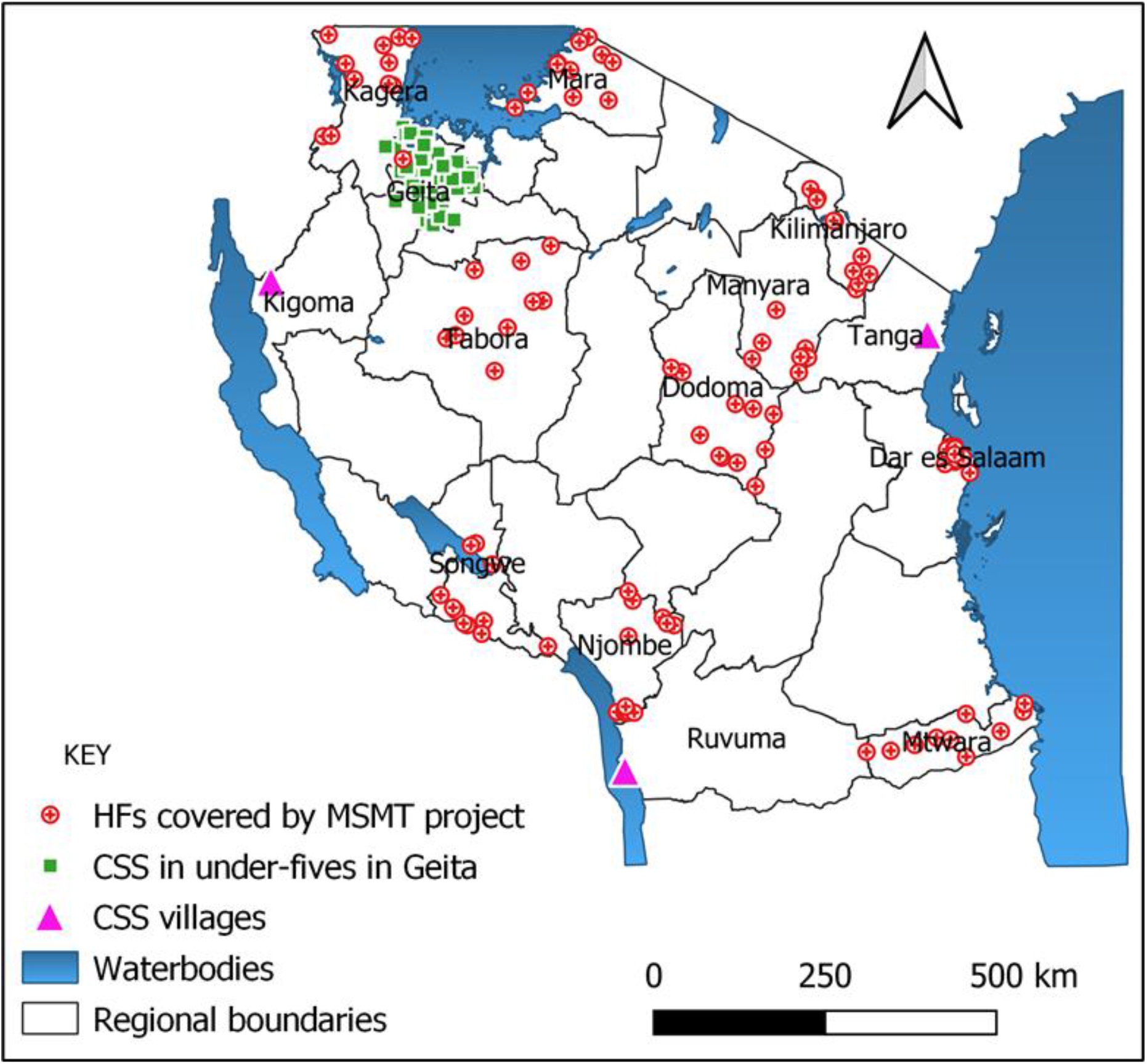
Map of Tanzania showing regions that were involved in the MSMT surveys in 2021. HFs = health facilities CSS = community cross-sectional surveys

### Enrollment of study participants and sample collection procedures

For each participant, finger prick blood was collected for detection of malaria infection by RDTs and preparation of dried blood spots (DBS) on Whatman 3 MM CHR filter papers (Cytiva, Marlborough, MA, USA) as described previously [29]. A total of 18,526 DBS samples were collected in the 14 regions; and 8,425 (45.5%) were RDT positive and 10,101 (54.5%) were RDT negative. From these, 4,776 (25.8%) were randomly selected for molecular analysis by quantitative polymerase chain reaction (qPCR) including 2,685 (26.6%) RDT negative samples, which were analysed for subpatent *P. falciparum* infection (Fig. 2).

**Fig. 2.**
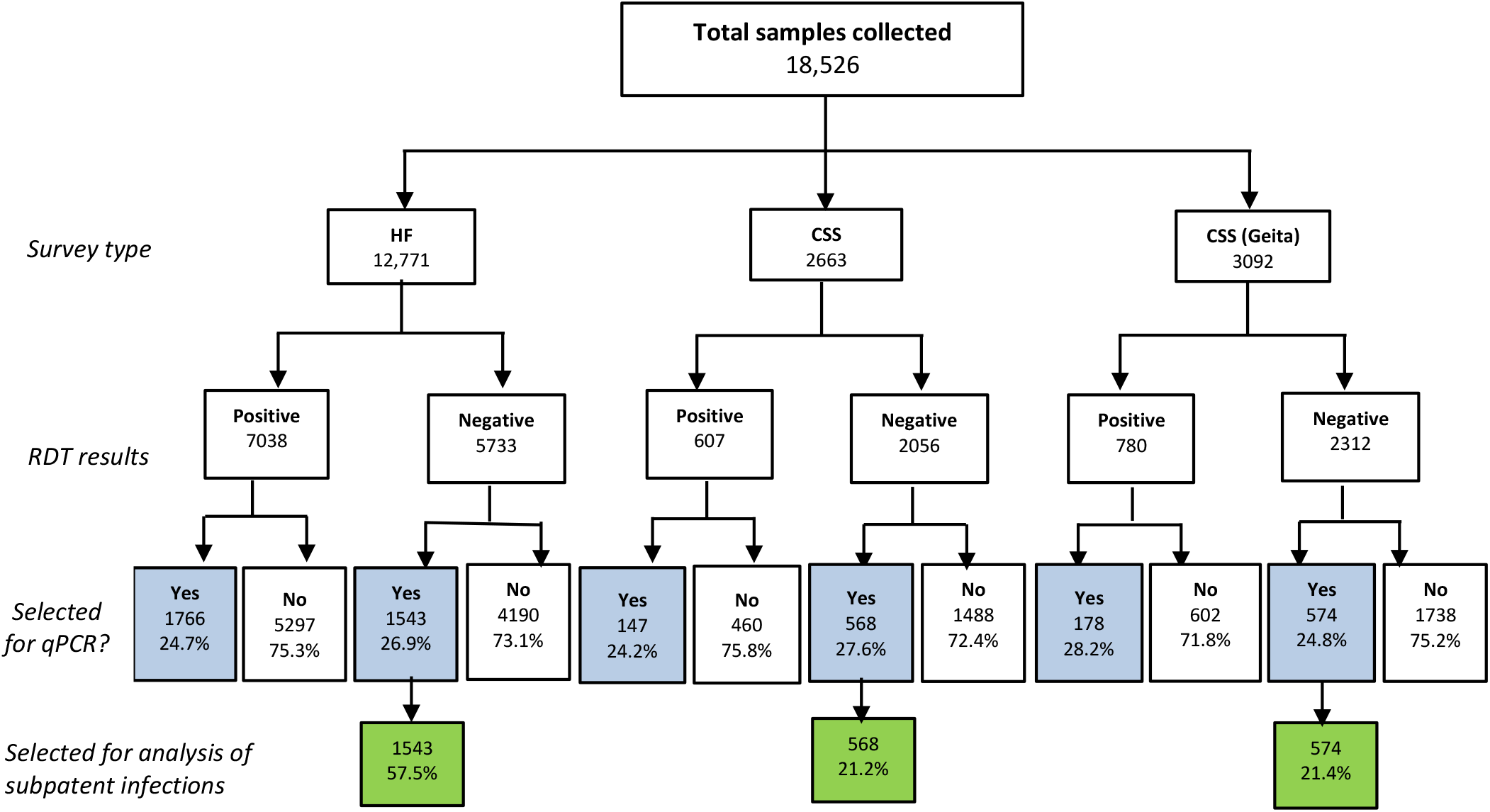
Flowchart showing how the samples used for this analysis were selected. **HF**=health facility **CSS**=community cross-sectional survey

**Fig. 3:**
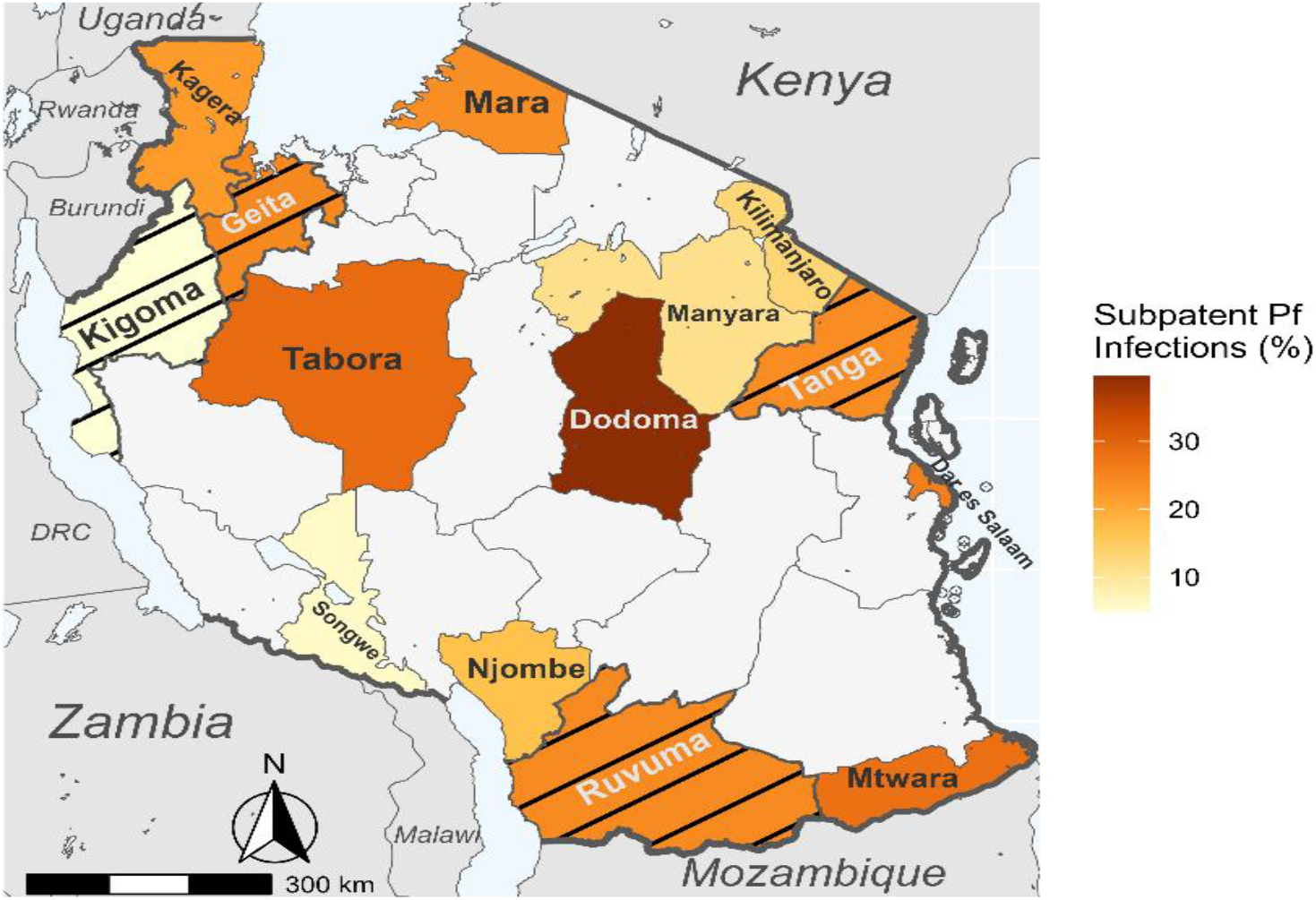
Map of Tanzania showing the proportion of symptomatic patients or asymptomatic individuals with subpatent infections in the studied regions. Black bars indicate regions where CSS surveys occurred, while those with solid fill indicate health facility surveys.

### Laboratory analysis

#### DNA extraction

Genomic DNA was extracted from DBS samples (three punches of 6 mm each per sample) using Tween-Chelex 100 (Bio-Rad Laboratories, <u>Hercules</u>, <u>C</u>A, USA) as previously described, with minor modifications [29,31]. Briefly, the punched DBS samples were incubated in 1 mL of 0.5% Tween-20 (Sigma) in phosphate buffered saline (PBS) (Thermo Fisher Scientific, USA) and placed on a shaker overnight at room temperature. After washing with 1x PBS and boiling at 95°C in Chelex 100 resin (Bio-Rad Laboratories, <u>Hercules, C</u>A, USA), a final volume of 150 μL of DNA was collected. After further centrifugation, OT-2 automated liquid handler (Opentrons Labworks, New York, USA) was used to make an aliquot of 50μL Chelex-free DNA. This aliquot was kept at −20°C until use in PCR assays.

#### Quantitative real-time PCR

Quantitative real-time PCR assay targeting the 18S ribosomal RNA (rRNA) was performed according to previously published methods [28,30,32]. Detection was done using TaqMan probe assay and parasitaemia quantification was based on standard curves generated using standard dilutions of plasmid DNA from MR4 (MRA-177, BEI Resources, Manassas, VA, USA) as previously described [28]. Parasitemia was estimated based on the assumption of six 18S rRNA gene copies per parasite genome [32] and then multiplied by four to account for the dilution of eluted DNA relative to the initial blood volume [33].

### Data management and analysis

Data from community surveys were collected using tools configured and installed on tablets, running Open Data Kit (ODK) software. The data were directly transmitted to a central data server located at the National Institute for Medical Research (NIMR) in Dar es Salaam, Tanzania. Health facility survey data were collected through paper questionnaires and double entered into a Microsoft® Access® LTSC MSO (Version 2405) database. All the data were transferred to a Microsoft® Excel® LTSC MSO (Version 2405), cleaned, checked for consistency and transferred to Stata version 13 (STATA Corp, Inc., 2015) for further cleaning and analysis. Descriptive statistics, including means, frequencies, and proportions were used to summarize the data. Chi-square tests were used to assess bivariate relationships between categorical variables and the prevalence of subpatent infections. Univariate and multivariate logistic regression analyses were used to identify factors associated with subpatent malaria infection status. Relationships between variables were presented as odds ratios (ORs) with 95% confidence intervals (CIs). All the factors with p-values <0.25 in the univariate analysis were included in the multivariate logistic regression model. Such variables were age group (under-fives, children aged 5-15 and individuals age>=15yrs), sex, fever status and transmission strata. Multivariate Linear regression analysis was used to assess the association between parasite density and the independent variables such as age group, sex, fever status, and transmission stratum. The assumption of normality for the parasite density distribution was tested using histograms and Shapiro test. As the parasite density was not normally distributed, it was log-transformed using the natural logarithmic function and analyzed to generate geometric mean parasite density with 95% CI. The parasite density in different strata was compared using Tukey’s honest significant difference (Tukey’s HSD) test. A p-value <0.05 was considered statistically significant. The regional-level map of positivity rate for subpatent infections was created using the R package *sf* (version 1.0–9) based on shape files available from GADM.org and naturalearthdata.com accessed via the R package *rnaturalearth* (version 0.3.2).

## Results

### Baseline characteristics of the study population

From among 4776 (25.8%) samples for which qPCR results were available, 2685 (56.2%) were negative by RDT and were used for this analysis (Figure 2). Among participants included in the analysis, age and sex information were available for 2040 (76.0%) and 2046 (76.2%) of participants, respectively. The median age (Interquartile range; IQR) of all participants was 8 years (4.4-25.7) and for the individuals selected for this study, the median age (IQR) was 8 years (5-31) (range: 6 months to 87 years). Nearly half (46.0%, n=1235/4776) of those included in this analysis were under-fives, 322 (12.0%) were school children (5-15 years old), and 1128 (42.0%) were adults (>15 years). The sex distribution was female-skewed, with 61.1% (n=1251/4776) female participants, reflecting the gender distribution in the main dataset where 56.1% (n=8651/18,526) were female (Table 1a). The majority (57.5%, n=1543/4,776) of participants of this study were drawn from health facility surveys which enrolled febrile/ symptomatic patients (Fig. 2, Table 1b).

**Table 1a:**
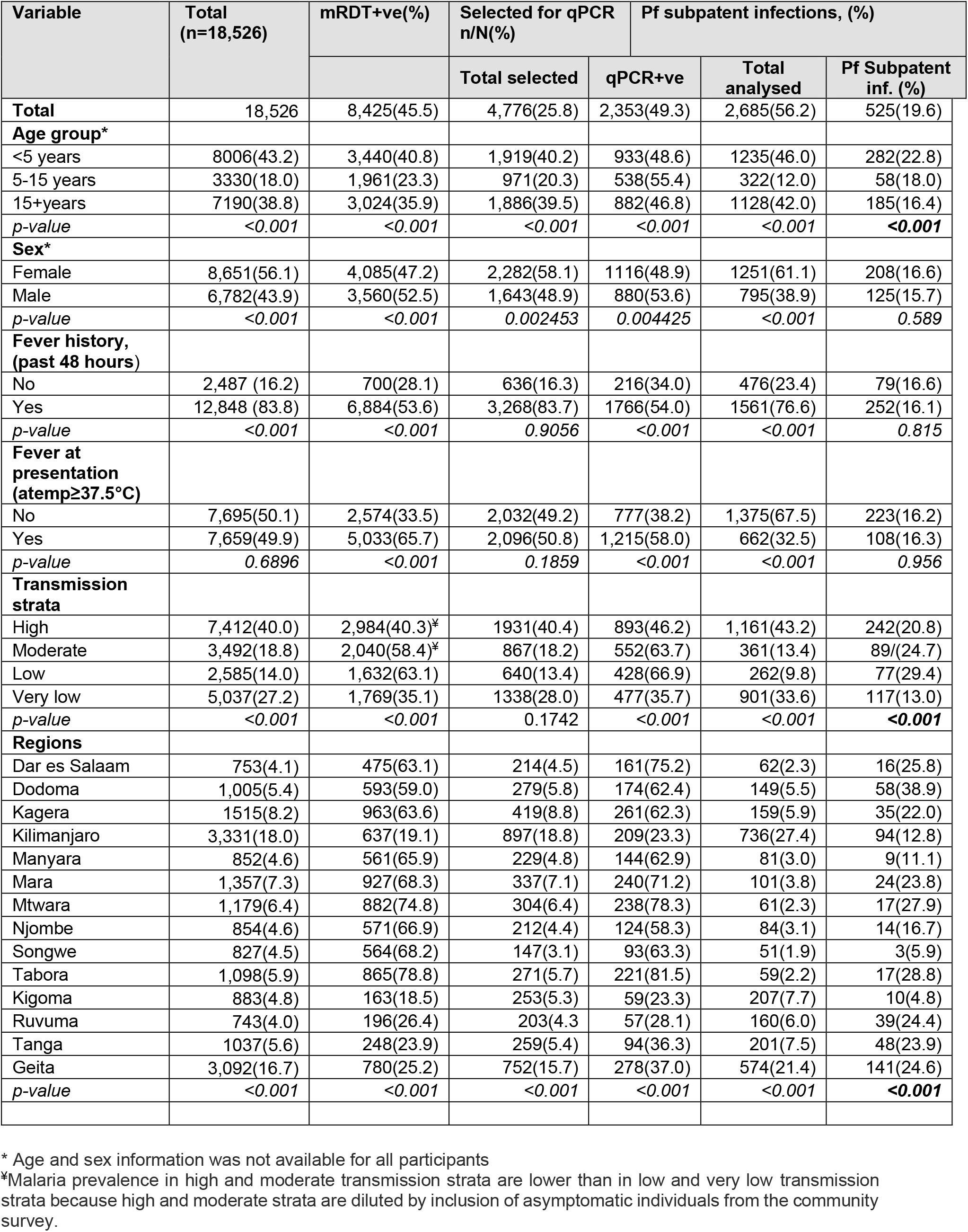
Demographic and clinical characteristics of individuals covered in the MSMT 2021 surveys and those selected for the analysis of subpatent infections.

| Variable | Total<br>(n=18,526) | mRDT+ve(%) | Selected for qPCR<br>n/N(%) |  | Pf subpatent infections, (%) |  |
| --- | --- | --- | --- | --- | --- | --- |
|  |  |  | Total selected | qPCR+ve | Total analysed | Pf Subpatent inf. (%) |
| **Total** | 18,526 | 8,425(45.5) | 4,776(25.8) | 2,353(49.3) | 2,685(56.2) | 525(19.6) |
| **Age group\*** |  |  |  |  |  |  |
| <5 years | 8006(43.2) | 3,440(40.8) | 1,919(40.2) | 933(48.6) | 1235(46.0) | 282(22.8) |
| 5-15 years | 3330(18.0) | 1,961(23.3) | 971(20.3) | 538(55.4) | 322(12.0) | 58(18.0) |
| 15+years | 7190(38.8) | 3,024(35.9) | 1,886(39.5) | 882(46.8) | 1128(42.0) | 185(16.4) |
| *p-value* | <0.001 | <0.001 | <0.001 | <0.001 | <0.001 | <0.001 |
| **Sex\*** |  |  |  |  |  |  |
| Female | 8,651(56.1) | 4,085(47.2) | 2,282(58.1) | 1116(48.9) | 1251(61.1) | 208(16.6) |
| Male | 6,782(43.9) | 3,560(52.5) | 1,643(48.9) | 880(53.6) | 795(38.9) | 125(15.7) |
| *p-value* | <0.001 | <0.001 | 0.002453 | 0.004425 | <0.001 | 0.589 |
| **Fever history,**<br>**(past 48 hours)** |  |  |  |  |  |  |
| No | 2,487 (16.2) | 700(28.1) | 636(16.3) | 216(34.0) | 476(23.4) | 79(16.6) |
| Yes | 12,848 (83.8) | 6,884(53.6) | 3,268(83.7) | 1766(54.0) | 1561(76.6) | 252(16.1) |
| *p-value* | <0.001 | <0.001 | 0.9056 | <0.001 | <0.001 | 0.815 |
| **Fever at presentation**<br>**(atemp≥37.5°C)** |  |  |  |  |  |  |
| No | 7,695(50.1) | 2,574(33.5) | 2,032(49.2) | 777(38.2) | 1,375(67.5) | 223(16.2) |
| Yes | 7,659(49.9) | 5,033(65.7) | 2,096(50.8) | 1,215(58.0) | 662(32.5) | 108(16.3) |
| *p-value* | 0.6896 | <0.001 | 0.1859 | <0.001 | <0.001 | 0.956 |
| **Transmission strata** |  |  |  |  |  |  |
| High | 7,412(40.0) | 2,984(40.3)‡ | 1931(40.4) | 893(46.2) | 1,161(43.2) | 242(20.8) |
| Moderate | 3,492(18.8) | 2,040(58.4)‡ | 867(18.2) | 552(63.7) | 361(13.4) | 89(24.7) |
| Low | 2,585(14.0) | 1,632(63.1) | 640(13.4) | 428(66.9) | 262(9.8) | 77(29.4) |
| Very low | 5,037(27.2) | 1,769(35.1) | 1338(28.0) | 477(35.7) | 901(33.6) | 117(13.0) |
| *p-value* | <0.001 | <0.001 | 0.1742 | <0.001 | <0.001 | <0.001 |
| **Regions** |  |  |  |  |  |  |
| Dar es Salaam | 753(4.1) | 475(63.1) | 214(4.5) | 161(75.2) | 62(2.3) | 16(25.8) |
| Dodoma | 1,005(5.4) | 593(59.0) | 279(5.8) | 174(62.4) | 149(5.5) | 58(38.9) |
| Kagera | 1515(8.2) | 963(63.6) | 419(8.8) | 261(62.3) | 159(5.9) | 35(22.0) |
| Kilimanjaro | 3,331(18.0) | 637(19.1) | 897(18.8) | 209(23.3) | 736(27.4) | 94(12.8) |
| Manyara | 852(4.6) | 561(65.9) | 229(4.8) | 144(62.9) | 81(3.0) | 9(11.1) |
| Mara | 1,357(7.3) | 927(68.3) | 337(7.1) | 240(71.2) | 101(3.8) | 24(23.8) |
| Mtwara | 1,179(6.4) | 882(74.8) | 304(6.4) | 238(78.3) | 61(2.3) | 17(27.9) |
| Njombe | 854(4.6) | 571(66.9) | 212(4.4) | 124(58.3) | 84(3.1) | 14(16.7) |
| Songwe | 827(4.5) | 564(68.2) | 147(3.1) | 93(63.3) | 51(1.9) | 3(5.9) |
| Tabora | 1,098(5.9) | 865(78.8) | 271(5.7) | 221(81.5) | 59(2.2) | 17(28.8) |
| Kigoma | 883(4.8) | 163(18.5) | 253(5.3) | 59(23.3) | 207(7.7) | 10(4.8) |
| Ruvuma | 743(4.0) | 196(26.4) | 203(4.3) | 57(28.1) | 160(6.0) | 39(24.4) |
| Tanga | 1037(5.6) | 248(23.9) | 259(5.4) | 94(36.3) | 201(7.5) | 48(23.9) |
| Geita | 3,092(16.7) | 780(25.2) | 752(15.7) | 278(37.0) | 574(21.4) | 141(24.6) |
| *p-value* | <0.001 | <0.001 | <0.001 | <0.001 | <0.001 | <0.001 |

\* Age and sex information was not available for all participants
‡Malaria prevalence in high and moderate transmission strata are lower than in low and very low transmission strata because high and moderate strata are diluted by inclusion of asymptomatic individuals from the community survey.

**Table 1b:** Demographic and clinical characteristics of individuals enrolled from health facilities.

| Variable | Total (n=12,771) | mRDT+ve(%) | Selected for qPCR n(%) |  | Pf subpatent infections (%) |  |
| --- | --- | --- | --- | --- | --- | --- |
|  |  |  | Total selected | qPCR+ve | Total analysed | Pf Subpatent infections |
| Total | 12,771 | 7,038 (55.1) | 3,309(25.9) | 1,865(56.4) | 1,543 (46.6) | 287(18.6) |
| **Age group\*** |  |  |  |  |  |  |
| <5 years | 4,459(34.9) | 2,576 (57.8) | 1,092(33.0) | 640(58.6) | 560 (36.3) | 133(23.8) |
| 5-15 years | 2,435(19.1) | 1,619 (66.5) | 728(22.0) | 455(62.5) | 196 (12.7) | 39(19.9) |
| 15+years | 5,877(46.0) | 2,843 (48.4) | 1,489(45.0) | 770(51.7) | 787 (51.0) | 115(14.6) |
| *p-value* | <0.001 | <0.001 | <0.001 | <0.001 | <0.001 | <0.001 |
| **Sex\*** |  |  |  |  |  |  |
| Female | 7,076 (55.4) | 3,763 (53.2) | 1,770(55.0) | 967(54.6) | 841 (56.7) | 136(16.2) |
| Male | 5,695 (44.6) | 3,275 (57.5) | 1,451(45.0) | 826(56.9) | 642 (43.3) | 103(16.0) |
| *p-value* | <0.001 | <0.001 | 0.5618 | 0.2048 | 0.06932 | 0.947 |
| **Fever history (past 48 hours)** |  |  |  |  |  |  |
| No | 1,130(8.9) | 612(54.2) | 272(8.5) | 144(52.9) | 132 (9.0) | 26(19.7) |
| Yes | 11,542(91.1) | 6,365(55.2) | 2,928(91.5) | 1,635(55.8) | 1342 (91.0) | 211(15.7) |
| *p-value* | <0.001 | 0.545 | 0.4 | 0.3916 | 0.4297 | 0.236 |
| **Fever at presentation (atemp≥37.5°C)** |  |  |  |  |  |  |
| No | 5,096(40.2) | 1,993(39.1) | 1,347(42.0) | 579(43.0) | 825 (56.0) | 129(15.6) |
| Yes | 7,595(59.8) | 5,007(65.9) | 1,863(58.8) | 1,210(64.9) | 649 (44.0) | 108(16.6) |
| *p-value* | <0.001 | <0.001 | 0.01653 | <0.001 | <0.001 | 0.602 |
| **Transmission strata** |  |  |  |  |  |  |
| High | 2,694(21.1) | 1,845 (68.4) | 723(21.8) | 499(69.1) | 220(14.3) | 52(23.6) |
| Moderate | 2,455(19.2) | 1,792(73.0) | 608(18.4) | 461(75.8) | 160(10.4) | 41(25.6) |
| Low | 2,585(20.2) | 1,632(63.1) | 640(19.3) | 428(66.9) | 262(17.0) | 77(29.4) |
| Very low | 5,037(39.4) | 1,769(35.1) | 1,338(40.4) | 477(35.7) | 901(58.4) | 117(13.0) |
| *p-value* | <0.001 | <0.001 | <0.001 | <0.001 | <0.001 | <0.001 |
| **Regions** |  |  |  |  |  |  |
| Dar es Salaam | 753 (5.9) | 475 (63.1) | 214 (6.5) | 161 (75.2) | 62 (4.0) | 16 (25.8) |
| Dodoma | 1,005 (7.9) | 593 (59.0) | 279 (8.4) | 174 (62.4) | 149 (10.0) | 58 (38.9) |
| Kagera | 1,515 (11.9) | 963 (63.6) | 419 (12.7) | 261 (62.3) | 159 (10.3) | 35 (22.0) |
| Kilimanjaro | 3,331 (26.1) | 637 (19.1) | 897 (27.1) | 209 (23.3) | 736 (47.7) | 94 (12.8) |
| Manyara | 852 (6.7) | 561 (65.8) | 229 (6.9) | 144 (62.9) | 81 (5.2) | 9 (11.1) |
| Mara | 1,357 (10.6) | 927 (68.3) | 337 (10.2) | 240 (71.2) | 101 (6.5) | 24 (23.8) |
| Mtwara | 1,179 (9.2) | 882 (74.8) | 304 (9.2) | 238 (78.3) | 61 (4.0) | 17 (27.9) |
| Njombe | 854 (6.7) | 571 (66.9) | 212 (6.4) | 124 (58.3) | 84 (5.4) | 14 (16.7) |
| Songwe | 827 (6.5) | 564 (68.2) | 147 (4.4) | 93 (63.3) | 51 (3.3) | 3 (5.9) |
| Tabora | 1,098 (8.6) | 865 (78.8) | 271 (8.2) | 221 (81.5) | 59 (3.8) | 17 (28.8) |
| *p-value* | <0.001 | <0.001 | <0.001 | <0.001 | <0.001 | <0.001 |

\* Age and sex information was not available for all participants

### Prevalence of subpatent infections

Among the 2685 RDT-negative samples tested, 525 (19.6%) were positive by qPCR (Table 1a).

#### Health facility survey

In the health facility survey, the prevalence was 18.6% overall (287/1543); prevalence was significantly higher in under-fives (23.8%, n = 133/560) compared to other age groups (p < 0.001). There were no significant differences by sex in prevalence of subpatent infections (16.2% in females vs 16.0% in males, unadjusted OR: 1.01, 95% CI 0.76 - 1.34; p = 0.947), though the prevalence of patent infections was significantly higher in males (57.5%) compared to females (53.2%; p < 0.001). The prevalence of sub-patent infection was also similar in individuals with or without fever at presentation or history of fever within 48 hours before the survey (p > 0.237 for both comparisons)(Table 1b). School children had a slightly, but not statistically significantly, higherodds of subpatent infections than children under 5 (aOR: 1.24, 95% CI 0.18 - 1.88; p = 0.307).

The prevalence was significantly lower in the very low transmission stratum (13.0%) compared to high transmission transmission strata (23.6%; aOR: 0.53, 95% CI 0.37 - 0.78; p < 0.001). The prevalence of subpatent infection was similar in low transmission to moderate and high transmission strata. The prevalence of subpatent infections was particularly high in Dodoma region (38.9%), making the overall prevalence highest in the low transmission stratum; 29.4%); sensitivity analysis excluding Dodoma region demonstrated that prevalence in low transmission areas dropped significantly to 16.8%. This was lower than high and moderate transmission areas (aOR: 0.65, 95% CI 0.36 - 1.16, p = 0.146).

#### Cross sectional survey

In the CSS group in the three regions (Kigoma, Ruvuma, and Tanga), the prevalence of subpatent infections was 17.1% (n = 97/568) and was significantly lower in under-fives (7.9%) compared to older individuals (>15.0%; p = 0.002), and in individuals from high compared to moderate transmission stratum (23.9 vs 15.5%; p = 0.01). The prevalence was similar among individuals of different sex (females 17.5%, males 14.4%; unadjusted OR: 1.27, 95% CI 0.76 - 2.13; p = 0.369) and those with or without fever at presentation () or history of fever within 48 hours before the survey (18.7% vs 15.4%; OR 1.26, 95% CI 0.81 - 1.98; p = 0.31) (Table 1c, Table 2). After adjusting for sex, transmission strata, fever status, and survey type, the risk of subpatent parasitemia in under-fives compared to older individuals remained significantly lower (aOR: 0.33, 95% CI 0.15 - 0.72, p = 0.005) (Table 2). The prevalence of subpatent infections was 24.6% (141/574) among under-fives in Geita region; significantly higher compared to that of under-fives in the other three CSS regions (7.9%; p < 0.001).

**Table 1c:** Demographic and clinical characteristics of individuals enrolled from community cross-sectional surveys.

| Variable | Total<br>(n=6,663) | mRDT+ve<br>(%) | Selected for qPCR (%) |  | Pf subpatent infections, (%) |  |
| --- | --- | --- | --- | --- | --- | --- |
|  |  |  | Total<br>selected | qPCR+ve | Total<br>analysed | Pf Subpatent<br>infections |
| **Total** | 2,663 | 607 (22.8) | 715 (26.8) | 210 (29.4) | 568 (79.4) | 97 (17.1) |
| **Age group** |  |  |  |  |  |  |
| <5 years | 455 (17.1) | 84 (18.5) | 97 (13.6) | 15 (15.5) | 79 (13.9) | 8 (7.9) |
| 5-15 years | 895 (33.6) | 342 (38.2) | 227 (31.7) | 83 (36.6) | 142 (25.0) | 19 (15.1) |
| 15+years | 1,313 (49.3) | 181 (13.8) | 391 (54.7) | 112 (28.6) | 347 (61.1) | 70 (20.5) |
| *p-value* | <0.001 | <0.001 | 0.00099 | 0.00061 | <0.001 | 0.002 |
| **Sex** |  |  |  |  |  |  |
| Female | 1,575 (59.2) | 322 (20.4) | 512 (72.7) | 149 (29.1) | 410 (72.8) | 72 (17.5) |
| Male | 1,087 (40.8) | 285 (26.2) | 192 (27.3) | 54 (28.1) | 153 (27.2) | 22 (14.4) |
| *p-value* | <0.001 | 0.00057 | <0.001 | 0.9 | 0.9923 | 0.86 |
| **History of fever**<br>**past 48 hours** |  |  |  |  |  |  |
| No | 1,357 (51.0) | 88 (6.5) | 364 (51.7) | 72 (19.8) | 344 (61.1) | 53 (15.4) |
| Yes | 1,306 (49.0) | 519 (39.7) | 340 (48.3) | 131 (38.5) | 219 (38.9) | 41 (18.7) |
| *p-value* | 0.1706 | <0.001 | 0.6757 | <0.001 | <0.001 | 0.332 |
| **Fever at**<br>**presentation**<br>**(atem>=37.5°C)** |  |  |  |  |  |  |
| No | 2,599 (97.6) | 581 (22.4) | 685 (95.8) | 198 (28.9) | 550 (96.8) | 94 (17.1) |
| Yes | 64 (2.4) | 26 (40.6) | 30 (4.2) | 5 (16.7) | 18 (3.2) | 0 (0.00) |
| *p-value* | <0.001 | 0.00099 | 0.0004374 | 0.2119 | 0.01385 | 0.84 |
| **Transmission**<br>**strata\*** |  |  |  |  |  |  |
| High | 1,626 (61.1) | 359 (22.1) | 456 (63.8) | 116 (25.4) | 367 (64.6) | 57 (15.5) |
| Moderate | 1,037 (38.9) | 248 (23.9) | 259 (36.2) | 94 (36.3) | 201 (35.4) | 48 (23.9) |
| *p-value* | <0.001 | 0.2918 | 0.08963 | 0.002905 | 0.4131 | 0.014 |
| **Regions** |  |  |  |  |  |  |
| Kigoma | 883 (33.2) | 163 (18.5) | 253 (35.4) | 59 (23.3) | 207 (36.4) | 18 (8.7) |
| Ruvuma | 743 (27.9) | 196 (26.4) | 203 (28.4) | 57 (28.1) | 160 (28.2) | 39 (24.4) |
| Tanga | 1,037 (38.9) | 248 (23.9) | 259 (36.2) | 94 (36.3) | 201 (35.4) | 48 (23.9) |
| *p-value* | <0.001 | 0.0004109 | 0.1828 | 0.004962 | 0.4825 | <0.001 |

\*The CSS group involves communities from high and moderate strata only because the survey was conducted conveniently and utilized an existing platform of communities for which census data is available.

**Table 2.** Logistic regression analysis to determine the odds of subpatent infections among RDT-negative samples from 14 regions of Mainland Tanzania.

| Characteristics | Health facilities survey |  | Community survey |  |
| --- | --- | --- | --- | --- |
|  | uOR, 95% CI, *p-value* | aOR, 95% CI, *p-value* | uOR, 95% CI, *p-value* | aOR, 95% CI, *p-value* |
| **Transmission strata** |  |  |  |  |
| High | Reference | Reference | Reference | Reference |
| Moderate | 1.11 (0.69 - 1.78), 0.66 | 1.10 (0.69 - 1.77), 0.69 | 2.04 (1.31- 3.17), 0.002 | 2.05 (1.31 - 3.21), 0.002 |
| Low | 1.34 (0.89 - 2.02), 0.16 | 1.35 (0.89 - 2.04), 0.148 |  |  |
| Very low | 0.48 (0.33 - 0.70), <0.001 | 0.53 (0.37 - 0.78), <0.001 |  |  |
| **Sex** |  |  |  |  |
| Male | Reference |  | Reference |  |
| Female | 1.01 (0.76- 1.34), 0.95 |  | 1.27 (0.76 - 2.13), 0.37 |  |
| **History of fever (past 48 hours)** |  |  |  |  |
| No | Reference |  | Reference |  |
| Yes | 0.76 (0.48- 1.20), 0.24 |  | 1.26 (0.81 - 1.98), 0.31 |  |
| **Fever at presentation (atemp ≥ 37.5°C)** |  |  |  |  |
| No | Reference |  |  |  |
| Yes | 1.08 (0.81- 1.42), 0.60 |  |  |  |
| **Age group** |  |  |  |  |
| <5 years | 1.82 (1.38 - 2.4), <0.001 | 1.40 (1.04 - 1.88), 0.03 | 0.33 (0.15 - 0.72), 0.005 | 0.33 (0.15 - 0.72), 0.005 |
| 5-14 years | 1.45 (0.97 - 2.17), 0.07 | 1.29 (0.85 - 1.94), 0.23 | 0.69 (0.39 - 1.20), 0.19 | 0.67 (0.38 - 1.18), 0.16 |
| 15+years | Reference | Reference | Reference | Reference |

atemp = xillary temperature; aOR =adjusted odds ratio; uOR = unadjusted odds ratio

### Parasite density

Parasite densities (geometric mean parasites per microlitre of blood (p/µL)) for subpatent infections varied significantly between regions (p < 0.001). The log-transformed median parasite density (IQR) was 6.9 (5.8 - 8.5) p/µL. Overall, parasite density was with significantly higher in the low compared to very low transmission stratum (11.4 vs 7.0 parasites/µL, p<0.001). The parasite density was highest in samples from Dodoma region (log_10_ 12.4 (8.8-16.3) p/µL) and lowest in Njombe (5.0, IQR = 4.5-5.7p/µL) (Fig. 4). Linear regression analysis revealed similar parasite densities between males and females in the health facility survey samples (males had 0.51 p/µL less than females, 95% CI −1.13 - 0.11, p = 0.11) while among those sampled in CSS, males (8.62 p/µL) had a statistically higher average parasite density than females (7.18 p/µL) (Adj Coef β 1.38 p/µL (95% CI 0.06 - 2.70, p = 0.04), though from a clinical standpoint this is not different. In the CSS, the parasite density increased in school children (aged 5 - 15 years) compared to those >15 years (8.7 p/µL vs 7.33 p/µL; Adj. Coef β 1.42, 95% CI 0.13-2.70, p=0.03), and increased non-significantly in under five children from health facility surveys (Table. 3). Tukey analysis of CSS parasitemia data revealed higher but non-significant mean parasite density in school children, and individuals from high (8.0 p/µL) as compared to moderate (7.1 p/µL) transmission strata, while in the health facility surveys, the mean parasite density was significantly higher in under-fives (8.1 p/µL) than in older age groups (6.1 p/µL for school children and 7.0 p/µL for adults), and in individuals from low (11.4 p/µL) versus high (6.7 p/µL) transmission strata (Fig. 5).

**Fig. 4.**
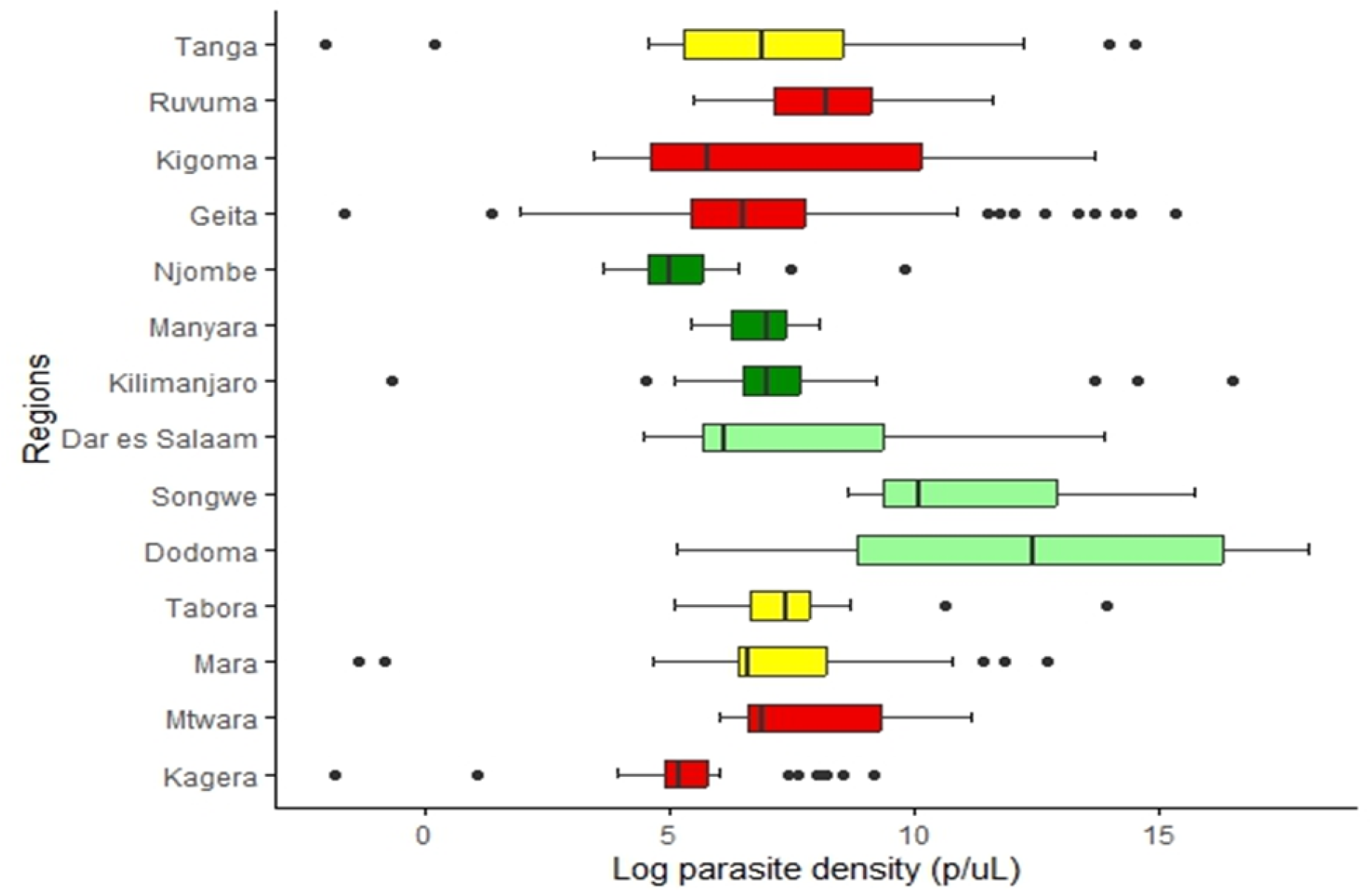
Median parasite densities in the different regions with varying transmission intensities. Transmission strata are classified as high (red), moderate (yellow), low (light green), and very low (green). The four top regions (Tanga, Ruvuma, Kigoma, and Geita) were involved in CSS while the rest were involved in health facility surveys.

**Fig. 5.**
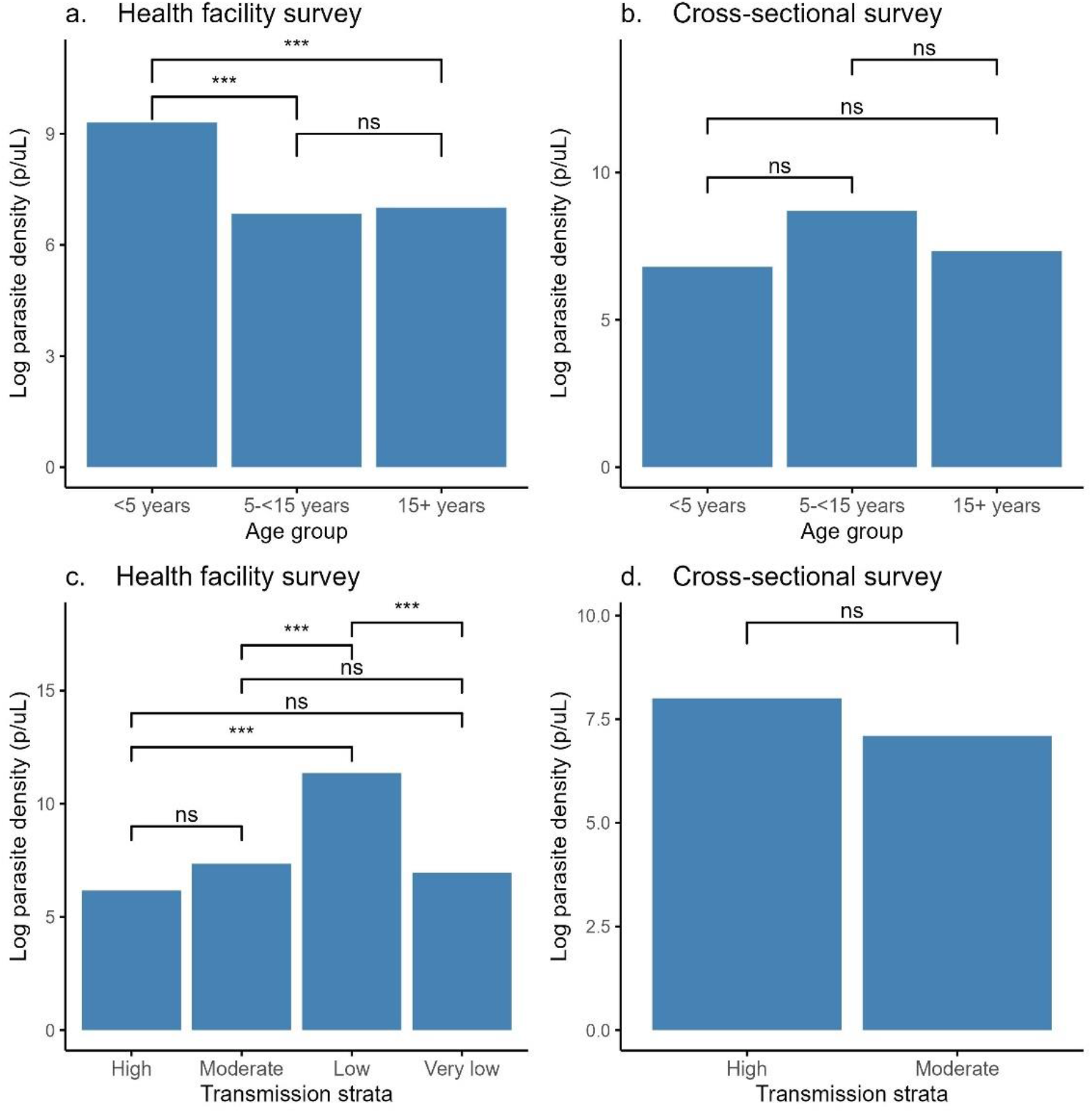
Tukey analysis of subpatent parasite density by age groups (a[health facility survey]; b[community survey]) and transmission Strata (c[health facility survey]; d[community survey]). Level of significance is shown as follows: ** p<0.01, *** p<0.001, ns=no significant difference.

**Table 3.** Results of linear regression analysis of factors associated with parasite density of subpatent infections among RDT-negative samples from 14 regions of Mainland Tanzania.

|  | Health facility model |  |  |  | Cross-sectional survey model |  |  |  |
| --- | --- | --- | --- | --- | --- | --- | --- | --- |
| Variable | Crude<br>Coef $\beta$ | 95% CI, P-value | Adj<br>Coef $\beta$ | 95% CI, P-value | Crude<br>Coef $\beta$ | 95% CI, P-value | Adj<br>Coef $\beta$ | 95% CI, P-value |
| Sex |  |  |  |  |  |  |  |  |
| Male | -0.431 | -1.052 - 0.191, p=0.173 | -0.51 | -1.128 - 0.109, p = 0.106 | 1.437 | 0.2 00 - 2.674, p = 0.023 | 1.382 | 0.061-2.703, p = 0.041* |
| Female |  | Reference |  | Reference |  | Reference |  | Reference |
| Age in years | -0.009 | -0.025 - 0.007, p=0.284 |  |  | -0.025 | -0.049 - - 0.002, p = 0.04 |  |  |
| Age group |  |  |  |  |  |  |  |  |
| <5 years | 2.297 | 1.464 - 3.131, p=0.003 | 0.194 | -0.494 - 0.882, p = 0.579 | -0.533 | -2.452 - 1.386, p = 0.583 | -1.54 | -3.859 - 0.78, p = 0.191 |
| 5-15 years | -0.172 | -1.385 - 1.041, p=0.78 | -0.269 | -1.153 - 0.616, p = 0.55 | 1.374 | 0.043 - 2.704, p = 0.043 | 1.416 | 0.133 - 2.7, p = 0.031* |
| > 15 years |  | Reference |  | Reference |  | Reference |  | Reference |
| Transmission strata |  |  |  |  |  |  |  |  |
| Moderate | 1.189 | 0.005 - 2.373, p = 0.049 | 1.248 | 0.251 - 2.245, p = 0.014 | -0.905 | -1.954 - 0.145, p = 0.09 | -0.162 | -1.292 - 0.967, p = 0.776 |
| Low | 5.187 | 4.169 - 6.205, p = 0 | 2.276 | 1.181 - 3.372, p < 0.001 |  |  |  |  |
| Very low | 0.789 | -0.156 - 1.734, p = 0.102 | 0.759 | -0.039 - 1.558, p = 0.062 |  |  |  |  |
| High |  | Reference |  | Reference |  |  |  |  |
| Fever at presentation |  |  |  |  |  |  |  |  |
| Yes | -0.157 | -0.781 - 0.470, p = 0.622 | -0.378 | -0.999 - 0.244, p=0.232 |  |  |  |  |
| No |  | Reference |  | Reference |  |  |  |  |

40 atemp = axillary temperature; aOR =adjusted odds ratio; uOR = unadjusted odds ratio

## Discussion

Subpatent malaria infections are potential reservoirs for persistent malaria transmission, thus, they are a threat to the ongoing malaria elimination plans in malaria-endemic countries [14,16], particularly in elimination or pre-elimination areas [14,26,34–37]. Due to this threat, in areas nearing elimination programmes must ensure that all cases, including subpatent infections, are tracked and treated. Particularly in low transmission areas, where the prevalence of subpatent infections was the highest, this requires more sensitive diagnostic tools [34,38,39].

The present study assessed subpatent infections among symptomatic patients from health facilities and asymptomatic individuals from communities and demonstrated that the prevalence of subpatent *P. falciparum* infections is high, at 19.6% overall, with marked heterogeneity among the studied regions. The positivity rates were higher in regions with low transmission (similar to moderate and high transmission areas) and then fell in those with very low transmission. The heterogeneity in subpatent infections reflects an inverse relationship to the patent malaria infections which has been reported in areas of different endemicity in Mainland Tanzania [4–6] and elsewhere [16,26,40]. This heterogeneity in patent infections is attributed to scaled-up interventions which have resulted in a shift of malaria epidemiology, with some areas transitioning from hyper-endemic to hypo-endemic transmission [4,41]. The low RDT positivity rates observed in high and moderate transmission strata in this study are a result of inclusion of the community cross-sectional survey data, most of which came from asymptomatic individuals and only included communities from high and moderate strata. The high prevalence of subpatent infections in areas of low transmission highlights the urgent need to design and implement more efforts to eliminate malaria infections. The prevalence of subpatent infections was particularly high in Dodoma region; sensitivity analysis excluding Dodoma suggested that this region may be driving the high likelihood of subpatent infections in low transmission areas. When Dodoma was excluded, there was a low likelihood of subpatent infections in low compared to high and moderate transmission areas. Thus, future studies are needed to monitor these trends and confirm the pattern seen in this study.

Subpatent infections were less prevalent in asymptomatic under-fives than in older age groups. This is most likely because this group has had the least exposure to malaria and thus have not yet developed naturally acquired immunity to suppress the infection, so most infections are patent and cause clinical symptoms [42]. Many studies have reported a higher prevalence of subpatent infections in adults than in children, as well as in older children compared to younger children [16,17,19,26]. School children exhibit a high prevalence of patent infections, most of which are asymptomatic due to developing immunity [43–46]. Adults are more likely to have subpatent infections, as increasing age is associated with an increase in naturally acquired immunity, which suppresses the parasites to low densities that may be undetectable by routine tests [42].

This study did not find any significant association between fever and the prevalence of subpatent infections in participants recruited from health facilities, underscoring that these infections often remain asymptomatic, as has been reported from Malawi [47,48]. The lack of a significant association could also be related to the fact that the majority of individuals recruited from health facilities had a history of fever or had fever at the time of sample collection. In contrast, in community surveys, where most people were asymptomatic, submicroscopic parasitaemia was associated with fever. This was also reported in a cohort study in Uganda [49], indicating that subpatent infections may cause fever, although it is also possible that the fever was due to other concomitant infections. More studies to explore the contribution of subpatent malaria infection to fever are recommended.

The prevalence of subpatent infections increased with decreasing transmission intensity, but dropped again as transmission intensity became very low. This finding is in line with what has been reported previously [16,26], although other studies have reported a low prevalence of subpatent infections in low-transmission settings [50]. One possible reason could be that in high-transmission areas, the greater exposure to infectious bites results in more people with higher density infections compared to low transmission areas. These are then more likely to be detected and treated, clearing the infection [35].

In this study, the lowest prevalence of subpatent infections was found in very low transmission areas, such as Kilimanjaro, Manyara, and Njombe regions. In these areas, most people have likely lost any naturally acquired immunity to malaria they might have had; thus, the majority of infections become symptomatic and are treated [42]. In order to achieve the elimination goals stated in the 2021-2025 strategic plan, which proposes transitioning to malaria elimination in phases, the National Malaria Control Programme now implements malaria case-based surveillance pilots in areas with very low malaria burden, starting with three northern regions of Arusha, Kilimanjaro, and Manyara, with a plan to scale up to other very low malaria burden regions [4,51]. While rates of subpatent infections were lowest in these areas, subpatent infections were still detected in 13.0% of the tested individuals, which is high compared to what was reported in pre- and elimination areas in other countries where subpatent infection rates ranged between zero and five percent [18,37,48,52– 54]. Apart from gauging transition criteria based on patent infection prevalence and incidence, it may be necessary to also consider approaches targeting subpatent infections with the aim of reducing them to a level that does not pose a transmission threat in elimination settings. Therefore, more sensitive tests should be considered to improve the detection of subpatent infections.

The parasite densities were generally low and appeared to be heterogeneous across different regions and transmission strata. In this study, the highest average parasite density was found in the low transmission strata. This is in line with findings of other studies reporting that subpatent parasite density was negatively associated with transmission intensity [19,55,56]. The high parasite density in young children could be explained by their less developed immunity and could also indicate parasites transitioning from subpatent to patent status over time. Younger children are at higher risk of malaria infection, and the high density of subpatent infections, even in areas of low transmission, calls for continued attention to this group. There was a negative association between parasite density and fever, with febrile individuals having low-density subpatent infections. This is partially attributed to the fact that patients with patent parasitemia were not included in the modeling, and likely reflects that these individuals had an alternative etiology for their fever. Nonetheless, in the context of malaria control and elimination, even low-density infections can be infectious to mosquitoes, maintaining transmission in the population, and are thus important to identify and treat [14,37,57]. While the vast majority of subpatent infections are due to low density parasitemia; in situations where HRP2-based RDTs are used for parasite detection, deletion of the histidine-rich protein 2 and histidine-rich protein 3 genes (*pfhrp2* and *pfhrp3*) has also been implicated as a reason for the failure of RDTs to detect *P. falciparum* even with higher density infections [58–60]. It is important to continue monitoring for the occurrence and spread of hrp2/3 gene deleted parasites.

This study was part of a main study designed to maximize collection of RDT-positive samples. This may have an effect on representativeness of transmission strata since even individuals recruited from regions with low transmission may have come from areas with relatively higher transmission compared to the rest of the region. Future studies should consider random selection of health facilities to address this selection bias.

## Conclusion

Subpatent infections are common and heterogeneous in most parts of mainland Tanzania. Although regions with very low transmission had the lowest prevalence of subpatent infections, the prevalence is still higher than would be expected for an area targeting elimination. To manage subpatent infectious reservoirs in regions with very low levels of *P. falciparum* transmission, elimination programmes may consider adopting more sensitive detection methods for the case-based management (test-and-treat) approach or mass drug administration that does not rely on the sensitivity of diagnostic tests.

### Disclaimer

The findings and conclusions in this report are those of the authors and do not necessarily represent the official position of the U.S. Centers for Disease Control and Prevention.

## Data Availability

All data produced in the present study are available upon reasonable request to the authors

## List of abbreviations

aOR: adjusted odds ratio
CA: California
CDC: Centers for Disease Control and Prevention
CI: confidence interval
CSS: cross-sectional survey
DBS: dried blood spot
DNA: deoxyribonucleic acid
HF: health facility
IQR: interquartile range
ITNs: insecticide-treated nets
MA: Massachusetts
MR4: malaria research and reference reagent resource
MRCC: Medical Research Coordinating Committee
NIMR: National Institute for Medical Research
NMCP: National Malaria Control Programme
NMSP: National Malaria Strategic Plan
OR: odds ratio
PBS: Phosphate-buffered saline
PCR: polymerase chain reaction
qPCR: quantitative polymerase chain reaction
RDT: Rapid diagnostic test
sSA: sub-Saharan Africa
USA: United States of America
USAID: United States Agency for International Development
VA: Virginia
WHO: World Health Organization

## Ethics approval and consent to participate

This study is part of the MSMT project, whose protocol was submitted to, reviewed and approved by the Medical Research Coordinating Committee (MRCC) of NIMR, Tanzania (NIMR/HQ/R.8a/Vol.IX/3579). Research participants were asked and provided individual consent (or assent for children aged 7-17 years) for their participation in the survey and biobanking for future research. For children under the legal age of adulthood in Tanzania (<18 years), consent was obtained from a parent or guardian. An informed consent form was developed in English, translated into Kiswahili and used to obtain consent both verbally and in writing from all participants. All participants agreed and signed the consent or assent form or provided a thumbprint in conjunction with the signature of an independent witness in case the study participant was illiterate.

## Consent for publication

Not applicable

## Availability of data and materials

The datasets used and/or analysed during the current study are available from the corresponding author on reasonable request

## Authors’ contributions

MDS, JAB, JJJ, and DSI conceived the study. MDS, RAM, RB, CB, BML, RM, RB, and DP collected samples, extracted DNA, and performed qPCR analysis. CIM, JRG, JAB, JJJ, and DSI oversaw the project. DM, SA, AL, and SL facilitated data collection. MDS, FF, DJG, and ZRPH performed epidemiological and statistical analyses. ZRPH also generated the maps. MDS wrote the manuscript. JAB, JJJ, JRG, ZRPH, and DSI edited the manuscript. All authors read, contributed to, and approved the final manuscript.

## Acknowledgements

The authors wish to thank the participants and parents/guardians of all the children who participated in the surveys. We acknowledge the contributions of the following project staff and other colleagues who participated in the data collection and/or laboratory processing of the samples: Raymond Kitengeso, Ezekiel Malecela, Muhidin Kassim, Athanas Mhina, August Nyaki, Juma Tupa, Anangisye Malabeja, Emmanuel Kessy, George Gesase, Tumaini Kamna, Grace Kanyankole, Oswald Oscar, Richard Makono, Ildephonce Mathias, Godbless Msaki, Rashid Mtumba, Gasper Lugela, Gineson Nkya, Daniel Challe, Richard Malisa, Sawaya Msangi, Ally Idrisa, Francis Chambo, Kusa Mchaina, Neema Barua, Christian Msokame, Rogers Msangi, Salome Simba, Hatibu Athumani, Mwanaidi Mtui, Rehema Mtibusa, Jumaa Akida, Ambele Lyatinga and Tilaus Gustav. The finance, administrative and logistic support team at NIMR: Christopher Masaka, Millen Meena, Beatrice Mwampeta, Gracia Sanga, Neema Manumbu, Halfan Mwanga, Arison Ekoni, Twalipo Mponzi, Pendael Nasary, Denis Byakuzana, Alfred Sezary, Emmanuel Mnzava, John Samwel, Daud Mjema, Seth Nguhu, Thomas Semdoe, Sadiki Yusuph, Alex Mwakibinga, Rodrick Ulomi and Andrea Kimboi. Management of the National Institute for Medical Research, National Malaria Control Program and President’s Office-Regional Administration and Local Government (regional administrative secretaries of the 14 regions and district officials, staff from all 100 health facilities, and community health workers from the 3 regions involved in community cross-sectional surveys. Technical and logistics support from the Bill and Melinda Gates Foundation team is highly appreciated.

